# MedEdMENTOR AI: Can artificial intelligence help medical education researchers select theoretical constructs?

**DOI:** 10.1101/2023.11.16.23298661

**Authors:** Gregory Ow, Adam Rodman, Geoffrey V. Stetson

## Abstract

**INTRODUCTION:** Medical education scholarship often lacks a strong theoretical underpinning, with this gap most often affecting early-career researchers and researchers in the Global South. Large language models (LLMs) have shown considerable promise to augment human writing and creativity in a variety of settings. In this study, we describe the development of MedEdMENTOR — an online platform for medical education research with a library of over 250 theories — and the development and evaluation of MedEdMENTOR AI, an LLM containing knowledge from MedEdMENTOR and the first AI mentor for medical education research.

**METHODS:** From a postpositivist paradigm, we evaluated MedEdMENTOR AI by testing it against 6 months of qualitative research published in 24 core medical educational journals. In a blinded fashion, we presented MedEdMENTOR AI with only the phenomenon of the qualitative study, and asked it to recommend 5 theories that could be used to study that phenomenon.

**RESULTS:** For 55% (29 of 53) of studies, MedEdMENTOR AI recommended the actual theoretical constructs chosen in the respective qualitative studies.

**CONCLUSIONS:** Our data is preliminary, but it suggests that MedEdMENTOR AI and other LLMs can be highly effective in guiding medical education scholars towards theories that may be applicable in their research. Further research is needed to assess performance on other tasks in medical education research.

## Introduction

Medical education scholarship frequently operates without the backbone of a strong theoretical framework — a gap most pronounced for clinicians making the leap to academic research.^1^ This issue is exacerbated by the concentration of research expertise in affluent institutions, predominantly in the Global North, which sidelines a variety of critical perspectives.^2^

In response to this inequity, September 2023 marked the launch of MedEdMENTOR, an online platform featuring MedEdMENTOR AI, the first AI mentor for medical education research. MedEdMENTOR additionally features a suite of tools: instructional primers introducing foundational concepts in the field, a network summarizing over 250 applicable theories, and the first dedicated medical education literature search engine. The platform’s accumulation of 15,000 page views and registration of over 550 users from 42 distinct countries in its first seven weeks speaks volumes about its global necessity.

Large language models (LLMs) such as ChatGPT (OpenAI) encode large amounts of information. Techniques like retrieval-augmented generation allow LLMs to access labeled databases for more accurate retrieval.^3^ The deployment process was greatly simplified by the release of OpenAI’s public release of customized GPTs in November 2023, enabling MedEdMENTOR’s founders to develop MedEdMENTOR AI. MedEdMENTOR AI contains knowledge from MedEdMENTOR’s instructional primers to help answer questions and guide users through the complexities of medical education scholarship. Early feedback from the user base has pointed to MedEdMENTOR AI’s effectiveness.

We sought to analyze MedEdMENTOR AI’s performance on a specific task: recommending theories that would reasonably frame a given phenomenon. This task is important because proper theoretical framing of education projects facilitates investigation of key aspects of problems and effective communication of findings across diverse learning contexts.

## METHODS

### Extracting phenomenon-theory pairs from the literature

Approaching this project from a postpositivist paradigm, we first built a dataset of phenomenon-theory pairs (see Table) using an innovative method of extracting the research phenomena and corresponding theories outlined in manuscripts (see Supplement).

**Table 1.** Selected phenomenon-theory pairs with answers from MedEdMENTOR AI.

| Research phenomenon | Actual Theory Used | MedEdMENTOR AI Answer |
| --- | --- | --- |
| The conceptualization and application of a framework that examines the interplay of various social identities and their positioning within power structures in the context of medical education. | Intersectionality | **Intersectionality Theory**<br>Critical Race Theory<br>Positionality Theory<br>Feminist Pedagogy<br>Transformative Learning Theory |
| The negotiation of tasks and competencies among healthcare students working together in an interprofessional team during clinical placements. | Communities of Practice | Interprofessional Education Framework<br>**Communities of Practice**<br>Zone of Proximal Development<br>Activity Theory<br>Team-Based Learning Theory |

First, we used PubMed to select the most recent 6 months of publications (June - November 2023) in the MEJ-24 that contained the word “theory.” The MEJ-24 is a set of 24 medical education journals suggested to form a “core” of medical education.^4^ Articles were excluded if they did not have an abstract available, if they were not related to medical education, if they were not research studies, or if they did not explicitly state the theory or phenomenon under investigation.

We then provided the resulting abstracts to GPT-4-1106 to extract the medical education research phenomena and the corresponding theories used (see Supplement). GPT-4-1106 was instructed to create blinded pairs, that is, to describe the phenomena under study without exposing the theories that were used and vice-versa. In order to validate this approach, we reviewed all abstracts by hand and found 0 errors in the accuracy of data extraction.

### LLM preparation

We compared 3 different LLMs:

#### (1) “Vanilla” GPT-4

The plain GPT-4-1106 accessed via API (temperature of 0).

#### (2) MedEdMENTOR AI

A customized OpenAI GPT that had access to 9 of MedEdMENTOR’s Lessons on theoretical frameworks and the importance of theory in medical education research. Additionally, this version had access to a document with examples of phenomena and their corresponding researcher-chosen theories which were extracted from an external set of abstracts (i.e. none of these were in the dataset used for evaluation).

#### (3) MedEdMENTOR AI with Ablation

The same as (2), however with the removal of the example document with phenomenon-theory pairs. This was used to investigate the relative importance of providing such a document.

To minimize variability, we used the same prompt for all LLMs, a simplified variation of a chain of density prompt (see Supplement) which instructs the LLM to provide a list of 5 relevant theories that “may help to clarify the underlying mechanisms pertaining to the phenomenon.”

### Evaluation scoring

Each LLM was provided with a series of research phenomena and instructed to generate 5 suggested theories. We then manually compared whether the LLM’s output list contained the actual theory that was used in the published study. LLM answers were marked as a match if the theory used was a direct synonym (e.g. “Kern’s Model” vs “Kern’s Model for Curriculum Development”) or if either theory was reasonably considered to be a subset of another (e.g. “self determination theory” and “intrinsic motivation” would be considered a match).

## RESULTS

114 articles were retrieved in total, published between June 2023 and November 2023. 61 articles were excluded leaving 53 abstracts.

1. “Vanilla” GPT-4 — 26 of 53 (49%) answers contained a match to the actual theory used in publication.
2. MedEdMENTOR AI — 29 of 53 (55%) answers contained a match to the actual theory used in publication.
3. MedEdMENTOR AI with Ablation — 24 of 53 (45%) answers contained a match to the actual theory used in publication.

## DISCUSSION

MedEdMENTOR AI shows promise in the task of selecting an appropriate theory for a medical education phenomenon. Furthermore, our ablation methodology suggests that the power of such tools will only grow as they are given more task-specific examples and further access to external knowledge.

The applicability of a theory to examine a specific phenomenon is a subjective determination that depends on a researcher’s preferred paradigm, identity, and life experiences, among many other things. Therefore, using the theory selected by the authors as the “gold standard” for evaluation of the LLM outputs is inherently a limitation of this study. Simply because the LLM did not suggest the theory that was actually chosen by the authors does not mean that the LLM is “wrong,” because there isn’t necessarily a “right.” Likewise, it is important that MedEdMENTOR AI provide a menu of theories to its users, like the five theories we prompted the LLMs to generate, such that they can each be deeply examined for a potential fit with their world view.

Using this evaluation methodology, the theories that can be tested against are limited to the set of theories that have already been used in MEJ-24 (i.e. in medical education). Further iterations of MedEdMENTOR AI will focus on differentiating between theories which have already been used in medical education and theories which would be novel to the field. Examining previously used theories can facilitate continued evolution of frameworks and their applications, and examining novel theories may be useful to further expand the bounds of medical education research.

## CONCLUSION

GPT-4 has already been shown to provide value in scientific discovery tasks.^5^ Our experience with MedEdMENTOR AI suggests that LLMs can have the same impact in the field of medical education research, augmenting the theoretical constructs of human researchers. These technologies will likely have an even greater impact on early-career researchers, community educators, and educators in the Global South. As educational research often focuses on thematic evaluations of large amounts of human-generated text, we suspect that LLMs are capable of assisting in far more than just selection of a theoretical construct, though additional research will be necessary.

## Data Availability

All data produced in the present study are available upon reasonable request to the authors.

https://mededmentor.org

## Supplement

### PubMed MEJ-24 Search Query

(“Academic medicine : journal of the Association of American Medical Colleges”[Jour] OR “Medical Education”[Jour] OR “Medical Teacher”[Jour] OR “Anatomical Sciences Education”[Jour] OR “BMC Medical Education”[Jour] OR “Advances in Health Sciences Education”[Jour] OR “Teaching and Learning in Medicine”[Jour] OR “Journal of Continuing Education in the Health Professions”[Jour] OR “Journal of Surgical Education”[Jour] OR “Journal of Graduate Medical Education”[Jour] OR “The Clinical Teacher”[Jour] OR “Medical Education Online”[Jour] OR “GMS Journal for Medical Education”[Jour] OR “Simulation in Healthcare”[Jour] OR “Advances in Medical Education and Practice”[Jour] OR “Education for Health”[Jour] OR “Perspectives on Medical Education”[Jour] OR “International Journal of Medical Education”[Jour] OR “Journal of Educational Evaluation for Health Professions”[Jour] OR “African Journal of Health Professions Education”[Jour] OR “Journal of Medical Education and Curricular Development”[Jour] OR “Canadian Medical Education Journal”[Jour]) + “theory”

### GPT-4 Extraction Prompt

You are an expert in medical education research. I will provide you with an abstract. Please read it silently. Then state the phenomenon being studied WITHOUT stating the education theory used. Then separately state the education theory used WITHOUT stating the phenomenon.

Phenomenon being studied:

Education theory used:

### GPT-4 Theory Suggestion Prompt

You are an expert in medical education research. I will provide you with a phenomenon. Please think deeply about this specific phenomenon, and give me nuanced education theories that may help to clarify the underlying mechanisms pertaining to the phenomenon. When I say nuanced, I mean to think of theories that apply to this phenomenon but not to other medical education research phenomena.

1. Return 5 nuanced and specific education theories.
2. Return 5 theories that are more nuanced and specific.
3. Return 5 theories that are even more nuanced and specific.

Do not explain your answers.

(4) Looking across your entire list, select the most directly applicable 5 theories.

